# Using Routine Ventilator Data to Visualize and Quantify Dynamic Compliance Trends Post-Surfactant Therapy in Preterm Infants: A Proof-of-Concept Study

**DOI:** 10.64898/2026.09.10.26362806

**Authors:** Hiroyuki Higashiyama

## Abstract

Using routinely recorded ventilator data from 16 preterm infants with respiratory distress syndrome, dynamic compliance trajectories after surfactant administration were smoothed with a moving-median filter and characterized with a four-parameter logistic model; 10 infants showed sigmoidal improvement, and among 11 infants who received a second surfactant dose, 6 showed subsequent improvement, suggesting that this approach can visualize and quantify individual compliance recovery as a complementary tool to conventional clinical indicators.

## Introduction

Respiratory distress syndrome (RDS) in preterm infants arises from pulmonary surfactant deficiency, resulting in alveolar collapse and reduced lung compliance. Exogenous surfactant replacement remains a cornerstone of management. Although non-invasive ventilation strategies have become the standard of care, many preterm infants with RDS still require mechanical ventilation [1].

Modern ventilators can calculate lung compliance on a breath-by-breath basis. Compared with adult patients, significant noise is inevitable in premature infants owing to their small size, rapid and active breathing mode, and the presence of airway leaks, which obscures the underlying physiological trajectory, limiting its bedside applicability. Reiss et al. reported a temporal trend in lung compliance during conventional ventilation in premature infants [2]. However, compliance was calculated manually and reported as a representative value (mean) for the study group, rather than on an individual basis. I hypothesized that a standardized approach to interpreting dynamic compliance (*C*_dyn_) time-series data could reveal the true physiological trend beneath this noise and evaluated a four-parameter logistic model for characterizing improvements in compliance.

## Methods

### Study Population

From April 2024 to July 2026, preterm infants born at ≤35 weeks gestation were screened for eligibility. Infants were included if they were diagnosed with RDS, received an initial dose of exogenous surfactant after admission to the neonatal intensive care unit, and required mechanical ventilation for more than 6 hours. Diagnosis of RDS was based on clinical presentation, stable microbubble rating, oxygen requirements, lung ultrasonography, and chest radiography [3].

Modified bovine minced surfactant (Surfacten®; Mitsubishi Tanabe Pharma, Osaka, Japan) was used. Infants were excluded if ventilator data were unavailable. Sixteen infants met the inclusion criteria. Eleven infants received a second dose of surfactant.

This retrospective study was approved by the Institutional Ethics Committee of Toyama Prefectural Central Hospital (approval ID: 68-58), which waived individual informed consent given the retrospective design; an opt-out approach allowed guardians to decline participation, in accordance with local ethical guidelines. No additional clinical interventions were performed.

### Data Collection

All infants were ventilated with a Babylog® VN600 ventilator (Dräger, Lübeck, Germany) in pressure control/assist control mode, with or without volume guarantee. Ventilator settings were adjusted to maintain an expiratory tidal volume of 5–6 ml/kg. Ventilator-derived parameters, including *C*_dyn_, were recorded continuously with time stamps and exported as comma-separated value files for analysis.

### Data Processing and Statistical Analysis

The initial surfactant administration was defined as time zero. *C*_dyn_ was normalized to body weight (_n_*C*_dyn_). Serial _n_*C*_dyn_ time-series plots were generated for each infant using data sampled every 5 min. A centered 7-point moving median filter was applied to address instability in real-time ventilator data. The moving median filter is a well-established method for noise reduction in physiological signals [4,5]. Missing periods of three or more consecutive observations were treated as interruptions in the time series, and smoothing was performed separately for each uninterrupted segment. Isolated missing values were ignored, and partial windows were used at segment boundaries. A window period of 35 min was selected based on visual inspection. All statistical analyses were performed using R version 4.6.1 (R Foundation for Statistical Computing, Vienna, Austria) within RStudio version 2026.6.0.242 (Posit Software, PBC, Boston, MA, USA).

### Logistic Model for C_dyn_ Progression

The temporal evolution of _n_*C*_dyn_ was modeled using a four-parameter logistic function implemented using the drc package in R [6]:

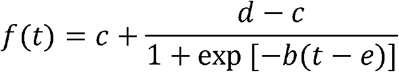

Here, b is the slope (maximal rate of compliance change); c, the lower asymptote (baseline compliance); d, the upper asymptote (plateau compliance); and e, the inflection point corresponding to the time of maximal rate of change (Figure 1A).

**Figure 1.**
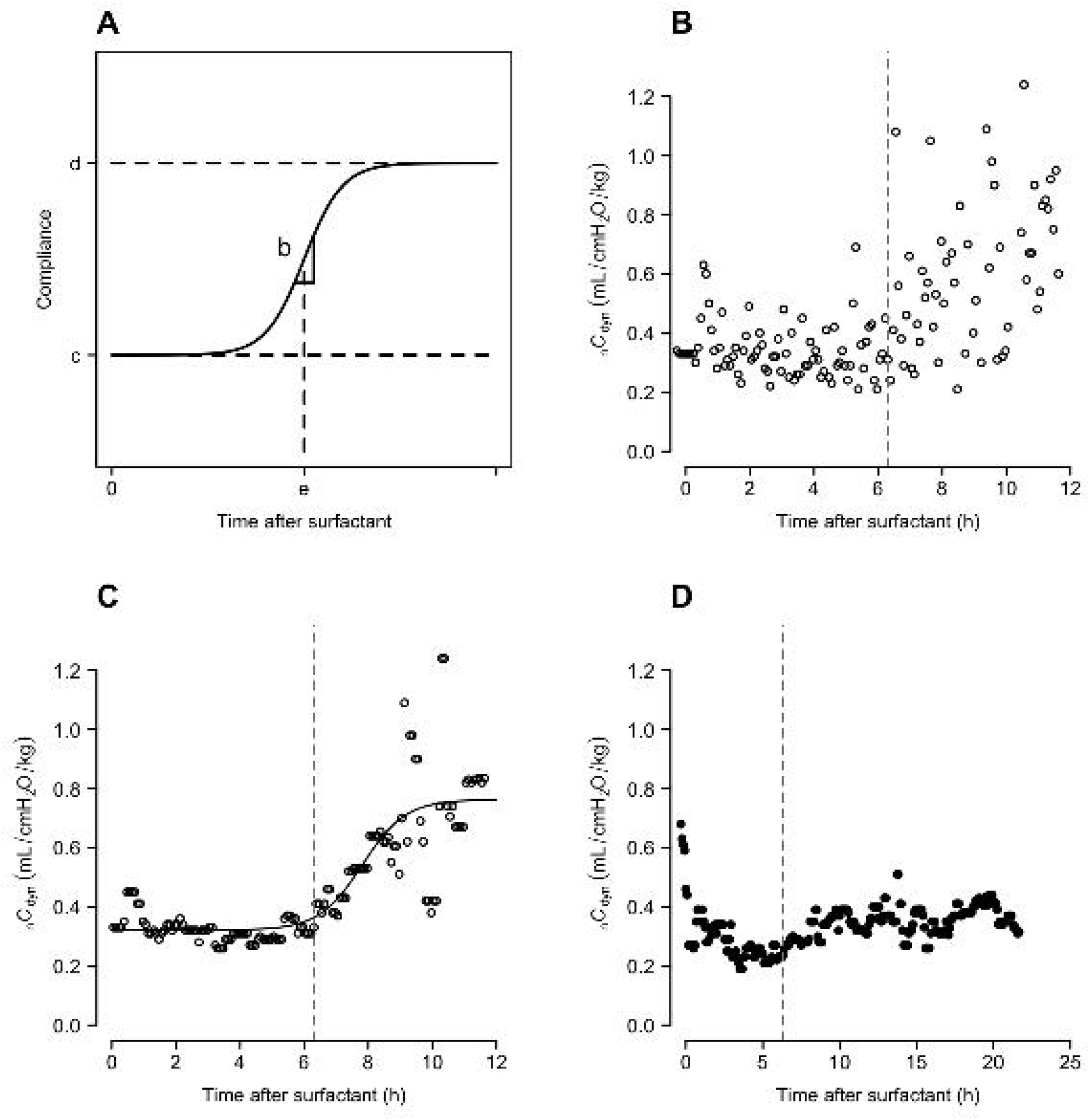
Normalized dynamic compliance trends. (A) Logistic curve and its parameters. (B) Representative sigmoidal _n_*C*_dyn_ trend in Case 1, with raw data. (C) Same data after moving median filtering, shown together with the fitted logistic curve. (D) Representative flat-pattern _n_*C*_dyn_ trend in Case 11 after moving median filtering. * The vertical dashed line indicates the timing of the second surfactant administration. C_dyn_, dynamic compliance; _n_C_dyn_, C_dyn_ normalized to body weight

## Results

### Patient Characteristics

Gestational age, growth status at birth, and the number of surfactant doses administered among the 16 infants are summarized in Table 1.

**Table 1.** Demographic data, goodness-of-fit metrics, and evaluated model parameters.

| Case | PMA <sup>1</sup> | Size <sup>2</sup> | Dose | Pattern | RSE <sup>3</sup> | R<sup>2</sup> <sup>4</sup> | b <sup>5</sup> | c <sup>5</sup> | d <sup>5</sup> | e <sup>5</sup> |
| --- | --- | --- | --- | --- | --- | --- | --- | --- | --- | --- |
| 1 | 35 | Appropriate | 2 | Sigmoid | 0.110 | 0.73 | 1.48 | 0.32 | 0.76 | 7.8 |
| 2 | 34 | Appropriate | 2 | Sigmoid | 0.128 | 0.69 | 0.80 | 0.30 | 0.74 | 9.4 |
| 3 | 33 | Appropriate | 1 | Sigmoid | 0.116 | 0.84 | 0.26 | 0.28 | 1.31 | 15.5 |
| 4 | 29 | Small | 2 | Sigmoid | 0.106 | 0.50 | 5.15 | 0.63 | 0.90 | 14.7 |
| 5 | 30 | Appropriate | 2 | Sigmoid | 0.041 | 0.89 | 0.18 | 0.16 | 0.64 | 7.0 |
| 6 | 30 | Small | 1 | Sigmoid | 0.127 | 0.53 | 1.64 | 0.57 | 0.87 | 8.4 |
| 7 | 29 | Appropriate | 1 | Sigmoid | 0.111 | 0.77 | 0.30 | 0.27 | 0.79 | 18.2 |
| 8 | 31 | Appropriate | 2 | Sigmoid | 0.064 | 0.74 | 0.93 | 0.32 | 0.56 | 15.3 |
| 9 | 32 | Appropriate | 1 | Sigmoid | 0.069 | 0.50 | 4.94 | 0.51 | 0.66 | 4.8 |
| 10 | 31 | Appropriate | 2 | Sigmoid | 0.128 | 0.79 | 0.95 | 0.25 | 0.84 | 5.7 |
| 11 | 34 | Appropriate | 2 | Flat | na | na | na | na | na | na |
| 12 | 29 | Appropriate | 2 | Flat | na | na | na | na | na | na |
| 13 | 29 | Appropriate | 2 | Flat | na | na | na | na | na | na |
| 14 | 31 | Appropriate | 1 | Flat | na | na | na | na | na | na |
| **15** | 32 | Appropriate | 2 | Flat | na | na | na | na | na | na |
| **16** | 32 | Appropriate | 2 | Flat | na | na | na | na | na | na |

<sup>1</sup> Post menstrual age (week)
<sup>2</sup> Size for gestational age
<sup>3</sup> Residual standard error
<sup>4</sup> Coefficient of determination
<sup>5</sup> Evaluated model parameters: b, slope (maximal rate of compliance change); c, lower asymptote (baseline compliance); d, upper asymptote (plateau compliance); e, inflection point corresponding to the time of maximal rate of change

### C_dyn_ Trend Patterns and Model Fit

The _n_*C*_dyn_ trajectory exhibited a sigmoid pattern in 10 of the 16 infants. A representative sigmoidal example (Case 1), comparing raw and moving median-filtered _n_*C*_dyn_ data with the fitted logistic curve, is shown in Figure 1B and 1C. A representative flat example (Case 11) is shown in Figure 1D.

### Residual Analysis

Residual histograms demonstrated approximately bell-shaped distributions centered near zero. Q–Q plots showed close alignment with the reference line between the −2 and +2 quantiles, supporting approximate normality (data not shown).

### Goodness of Fit and Parameter Estimates

Across the modeled cases, R^2^ values ranged from 0.50 to 0.89, and residual standard errors ranged from 0.041 to 0.128, indicating adequate model performance. The lower asymptote (c) ranged from 0.16 to 0.63 ml/cmH_2_O/kg, whereas the upper asymptote (d) ranged from 0.56 to 1.31 mL/cmH_2_O/kg. The slope (b) and inflection time (e) varied among infants, reflecting differences in the rate and timing of compliance improvement after surfactant therapy (Table 1).

## Discussion

This study demonstrates that a standardized, per-patient interpretation of *C*_dyn_ time-series data clearly delineates the trajectory of *C*_dyn_ following surfactant administration in preterm infants with RDS. This approach uses routinely recorded ventilator data, and the derived parameters can be calculated using commonly available spreadsheet software such as Microsoft Excel, making it readily accessible in clinical practice. The estimated logistic parameters may further serve as quantitative descriptors of treatment response, potentially enabling comparison across surfactant protocols.

Among all 11 cases that received a second surfactant dose, bedside assessment confirmed a persistently flat C_dyn_ trend before administration. The trend increased shortly after redosing in 6 of 11 cases, suggesting potential clinical utility for *C*_dyn_ trend monitoring when considering repeat dosing. A flat *C*_dyn_ trend despite initial surfactant therapy may serve as an objective indicator for re-treatment, although prospective validation remains necessary.

The primary limitation of this study is the small sample size, which limits generalizability. Additionally, the retrospective design precluded the assessment of associations between model-derived parameters and clinical outcomes. Larger prospective studies are needed to validate these findings and determine whether logistic model parameters can predict clinically relevant outcomes.

A *C*_dyn_ time series using data processed with a moving median provides clear, interpretable trend patterns after surfactant therapy in preterm infants with RDS. Logistic modeling of compliance trajectories yields quantitative parameters that may facilitate comparison of treatment responses across patients.

## Data Availability

All data produced in the present study are available upon reasonable request to the authors

## Acknowledgements

The author used ChatGPT (OpenAI) for English language editing to improve manuscript clarity and subsequently reviewed and edited the output as needed. Additionally, the manuscript was professionally edited for language and style by Editage. ChatGPT was also utilized to refine the R software codes, which were then thoroughly verified line by line by the author. The author takes full responsibility for the content of the published work.

## Data Availability Statement

The data supporting the findings of this study are available from the corresponding author upon reasonable request.

## Author Contributions

Hiroyuki Higashiyama: Conceptualization; Data curation; Formal analysis; Investigation; Methodology; Visualization; Writing – original draft; and Writing – review and editing.

## Abbreviations

RDS: respiratory distress syndrome
*C*_dyn_: dynamic compliance
_n_*C*_dyn_: *C*_dyn_ normalized to body weight

## References

1. Härtel C, Kribs A, Göpel W, Dargaville P, Herting E. Less invasive surfactant administration for preterm infants—state of the art. Neonatology. 2024;121(5):584–595. doi:10.1159/000540078

2. Reiss A, Lauth W, Wald M. Premature infants show consistently good lung compliance during conventional mechanical ventilation. Pediatr Pulmonol. 2025 Jan;60(1):e27419. doi:10.1002/ppul.27419

3. Hoshino Y, Futatani T, Kitamura S, et al. Lung ultrasound for predicting surfactant administration in ventilated and non-ventilated infants with respiratory distress syndrome: a multicenter prospective study. Eur J Pediatr. 2025 Nov;184(12):742. doi:10.1007/s00431-025-06580-0

4. An X, K Stylios GK. Comparison of motion artefact reduction methods and the implementation of adaptive motion artefact reduction in wearable electrocardiogram monitoring. Sensors (Basel). 2020 Mar;20(5):1468. doi:10.3390/s20051468

5. Honoré A, Forsberg D, Adolphson K, Chatterjee S, Jost K, Herlenius E. Vital sign-based detection of sepsis in neonates using machine learning. Acta Paediatr. 2023 Apr;112(4):686–696. doi:10.1111/apa.16660

6. Ritz C, Streibig JC. DRC: Analysis of Dose–Response Curves. R Package. doi:10.32614/CRAN.package.drc

